# Efficacy and Safety of Finerenone in Heart Failure With Preserved or Mildly Reduced Ejection Fraction: A Systematic Review and Meta-Analysis of Randomized Trials

**DOI:** 10.1101/2025.09.13.25335677

**Authors:** Rakhshanda Khan, Vamsi Pachchipulusu, Dharaneswari Hari Narayanan, Rahul Chidurala, Hanumanthu kavya tejaswini, Palak Patel, Raghu Vamsi Vanguru, Patel Yash Kalpeshbhai, Sowmiya Lakshmi, Nanditha Nandakishor, Harshawardhan Dhanraj ramteke, Abdus Sameey

## Abstract

**Introduction:** Heart failure with preserved (HFpEF) and mildly reduced ejection fraction (HFmrEF) accounts for nearly half of all heart failure cases, yet effective therapies remain limited. Finerenone, a novel non-steroidal mineralocorticoid receptor antagonist, offers anti-fibrotic and anti-inflammatory effects with improved safety compared to steroidal MRAs. We conducted a systematic review and meta-analysis to evaluate the efficacy and safety of finerenone in HFpEF/HFmrEF.

**Methods:** We systematically searched PubMed, Embase, Cochrane CENTRAL, Web of Science, and Scopus through September 2025, including only randomized controlled trials. Data extraction and risk of bias (RoB 2) assessments were performed independently by two reviewers. Pooled log risk ratios **log (RR)** and 95% confidence intervals (CIs) were calculated using random-effects models.

**Results:** Ten RCTs comprising 33,544 patients (20,279 males, 13,202 females) with a mean follow-up of 16.15 months were included. Finerenone significantly reduced major adverse cardiovascular events (MACE; log RR –0.11 [– 0.17, –0.05], p<0.001; I²=0%). All-cause mortality (log RR – 0.58, p=0.06), cardiovascular mortality (log RR –0.36, p=0.06), and rehospitalization (log RR – 0.66, p=0.07) trended favorably but were not statistically significant. Hyperkalemia risk was significantly increased (log RR 0.73, p<0.001).

**Conclusion:** Finerenone reduces MACE with an acceptable safety profile, albeit with higher hyperkalemia risk. These findings support finerenone as a promising therapy for HFpEF/HFmrEF, complementing existing treatment strategies.

## Introduction

Heart failure (HF) is a leading cause of morbidity and mortality worldwide, affecting more than 64 million people and placing a substantial burden on patients, caregivers, and healthcare systems [1]. While the pharmacologic management of heart failure with reduced ejection fraction (HFrEF) has been revolutionized by advances in guideline-directed medical therapy, therapeutic progress for patients with heart failure with preserved ejection fraction (HFpEF) and mildly reduced ejection fraction (HFmrEF) has been comparatively limited [2]. This disparity reflects both the heterogeneity of HFpEF pathophysiology and the lack of robust clinical trial evidence demonstrating consistent benefit of available therapies in this population.

HFpEF, defined by a left ventricular ejection fraction (LVEF) ≥50%, and HFmrEF, with LVEF 41–49%, account for nearly half of all HF cases [3]. Patients with HFpEF are typically older, more often female, and burdened by multiple comorbidities, including hypertension, diabetes, obesity, and atrial fibrillation [4]. These comorbidities contribute to systemic inflammation, endothelial dysfunction, and myocardial fibrosis—hallmarks of the syndrome that culminate in impaired diastolic function and increased filling pressures [5]. Despite its high prevalence, HFpEF has long been considered a “therapeutic graveyard” because most pharmacological interventions that are highly effective in HFrEF have failed to demonstrate consistent efficacy in HFpEF [6].

Recent guidelines by the American College of Cardiology (ACC), American Heart Association (AHA), and Heart Failure Society of America (HFSA) highlight this gap [7]. In the 2022 update, only SGLT2 inhibitors received a Class IIa recommendation for HFpEF/HFmrEF, based on the results of EMPEROR-Preserved and DELIVER, which demonstrated reduced risk of hospitalization for HF but no significant mortality benefit [8,9].

Diuretics remain a Class I recommendation for symptomatic relief, yet they do not modify long-term outcomes. Thus, the therapeutic landscape for HFpEF remains largely unsatisfactory, underscoring the need for novel, evidence-based treatment options.

Mineralocorticoid receptor antagonists (MRAs) represent one such therapeutic avenue. Aldosterone, through activation of the mineralocorticoid receptor, promotes myocardial hypertrophy, interstitial fibrosis, vascular stiffness, and systemic inflammation—pathways central to the pathophysiology of HFpEF [10]. Steroidal MRAs, such as spironolactone and eplerenone, have demonstrated benefit in HFrEF, but their role in HFpEF has been less clear. The TOPCAT trial, which investigated spironolactone in HFpEF, showed a reduction in HF hospitalizations but failed to demonstrate significant reductions in mortality or the composite primary outcome [11]. Moreover, the use of steroidal MRAs is often limited by safety concerns, particularly hyperkalemia and worsening kidney function.

Finerenone, a novel non-steroidal MRA, was developed to provide more selective receptor antagonism while minimizing off-target effects and metabolic adverse events. Unlike spironolactone and eplerenone, finerenone has a unique chemical structure that confers balanced tissue distribution and reduced risk of hyperkalemia [12]. Although finerenone was initially studied in patients with chronic kidney disease and type 2 diabetes—where the FIDELIO-DKD and FIGARO-DKD trials demonstrated significant reductions in cardiorenal outcomes [13,14]—its favorable pharmacologic profile and mechanistic rationale have positioned it as a promising candidate for HFpEF and HFmrEF populations.

The Finerenone in Heart Failure with Mildly Reduced or Preserved Ejection Fraction (FINEARTS-HF) trial was designed to directly address this question [15]. By enrolling patients with HFmrEF and HFpEF, irrespective of diabetes status, FINEARTS-HF explored the ability of finerenone to reduce clinically meaningful outcomes, including hospitalizations and cardiovascular events. Its results represent a pivotal advancement in the therapeutic landscape of HFpEF/HFmrEF and provide new insights into the role of mineralocorticoid receptor blockade beyond traditional agents.

The implications of these findings are considerable. Given the substantial unmet need for therapies that improve outcomes in HFpEF, finerenone may represent a paradigm shift in management. Its dual ability to target myocardial fibrosis and systemic inflammation, while offering an improved safety profile compared with conventional MRAs, could establish it as a cornerstone therapy for this challenging syndrome. Furthermore, as HFpEF frequently coexists with comorbidities such as diabetes and chronic kidney disease, the pleiotropic benefits of finerenone may extend beyond cardiac outcomes, contributing to holistic risk reduction in this highrisk population.

In this context, we conducted a systematic review and meta-analysis to evaluate the efficacy and safety of finerenone in patients with HFpEF and HFmrEF. By synthesizing evidence from recent clinical trials, particularly FINEARTS-HF, our study aims to clarify the therapeutic role of finerenone in this population and to assess its potential integration into guideline-directed management of HFpEF/HFmrEF. This analysis is timely and clinically relevant, as the field continues to search for effective, disease-modifying therapies in a population where evidence-based options remain scarce.

## Methods

### Literature Search

We systematically searched PubMed, Embase, Cochrane CENTRAL, Web of Science, and Scopus from inception to September 2025 to identify clinical trials evaluating **finerenone in heart failure with preserved (HFpEF) or mildly reduced ejection fraction (HFmrEF)**. Search terms combined “finerenone,” “nonsteroidal mineralocorticoid receptor antagonist,” “HFpEF,” and “HFmrEF,” with filters for randomized and controlled trials. Additional sources included **ClinicalTrials.gov**, major cardiology conference proceedings, and citation tracking. Two reviewers independently screened studies, extracted data, and assessed risk of bias (RoB 2) [16]. Discrepancies were resolved by consensus. The search adhered to PRISMA 2020 recommendations [17]. This Meta-analysis is registered on Prospero with the number CRD420251146338

### Study Selection, Data Extraction and Risk of Bias

All retrieved citations were de-duplicated and screened by two independent reviewers. Titles and abstracts were initially reviewed, followed by full-text evaluation based on predefined eligibility criteria (randomized controlled trials of finerenone in patients with HFpEF or HFmrEF). A standardized extraction form captured study design, population characteristics, interventions, comparators, primary and secondary outcomes, and follow-up duration. Discrepancies were resolved through consensus or adjudication by a third reviewer. Risk of bias was assessed using the **Cochrane RoB 2 tool**. Data extraction and evaluation adhered to **PRISMA 2020** guidelines to ensure methodological rigor and reproducibility.

### Statistical Analysis

We performed a meta-analysis using a random-effects model to account for between-study heterogeneity. Pooled treatment effects were expressed as log **risk ratios (**log RR**) with 95% confidence intervals (CIs)** for dichotomous outcomes and **mean differences (MDs) or standardized mean differences (SMDs)** for continuous outcomes. Heterogeneity was quantified with the **I² statistic** and Cochran’s Q test. Sensitivity analyses were conducted by sequential study exclusion. Publication bias was assessed using funnel plots and Egger’s regression when ≥10 studies were available. All analyses were performed with **Stata**, following **Cochrane Handbook** and **PRISMA 2020** recommendations.

## Results

### Demographics

A total of 625 studies were analysed out of which 10 were selected for meta-analysis [18–27] (Figure 1). A total of **33,481 patients** were included across eligible randomized trials. The population comprised **20,279 males** and **13,202 females**, with a mean follow-up duration of **16.15 months**. Among these, **17,470 patients** received finerenone, while **15,946 patients** were allocated to placebo or standard therapy. Baseline characteristics were well balanced between groups. The mean systolic blood pressure was slightly higher in the treatment arm (**132.6 ± 14.2 mmHg**) compared with controls (**127 ± 14 mmHg**). The overall mean body mass index of participants was **28.62 ± 2.5 kg/m²**, reflecting an overweight population consistent with comorbidities commonly observed in HFpEF and HFmrEF cohorts. Table S1.

**Figure 1.**
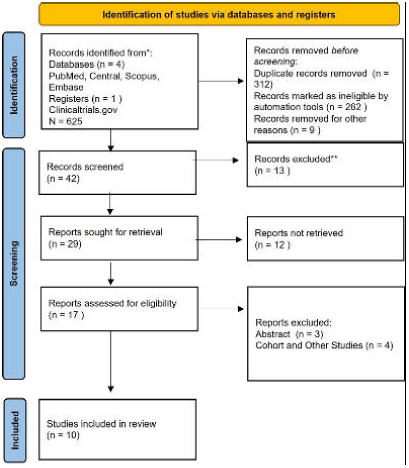
PRISMA Flow Diagram.

### All-Cause Mortality

The forest plot summarizes all-cause mortality risk ratios with finerenone versus control. Smaller early trials (Katayama, Sato, Pitt 2013) showed wide confidence intervals, reflecting low power. Filippatos and Bakris 2015 reported significant mortality reductions, while larger contemporary trials (Vardeny 2024, Bakris 2020, Pitt 2021, Solomon 2024, Butt 2025) demonstrated more precise estimates centered near null. Across major RCTs, effects were modest, with most confidence intervals crossing unity. The pooled analysis favored finerenone (log RR –0.58 [–1.19, 0.02]) but narrowly missed statistical significance (p=0.06). High heterogeneity (I²=98.4%) indicates variability across studies, warranting cautious interpretation. Figure 2.

**Figure 2.**
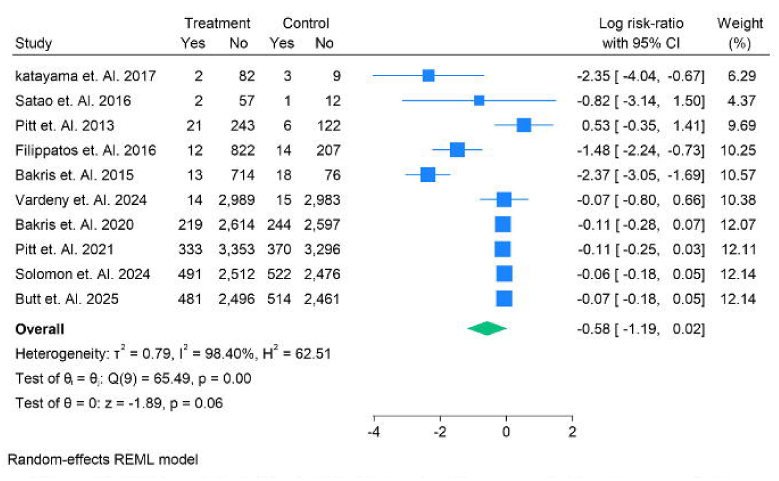
All Cause Mortality in Risk Ratios for Finerenone in the given population.

### Cardiovascular Mortality

This forest plot evaluates cardiovascular mortality with finerenone versus control. Early, small-scale studies (Katayama 2017, Sato 2016, Pitt 2013) suggested reductions but were limited by wide confidence intervals. Mid-sized trials (Filippatos 2016, Bakris 2015) showed mixed effects, with one favoring finerenone and the other trending neutral. Large contemporary trials (Bakris 2020, Pitt 2021, Solomon 2024, Butt 2025) consistently showed modest reductions in CV deaths, though effect sizes were small and confidence intervals overlapped unity. The pooled estimate (log RR –0.36, p=0.06) indicated a non-significant trend toward benefit.

Heterogeneity was high (I²=95.7%), reflecting variability among trials. Figure 3

**Figure 3.**
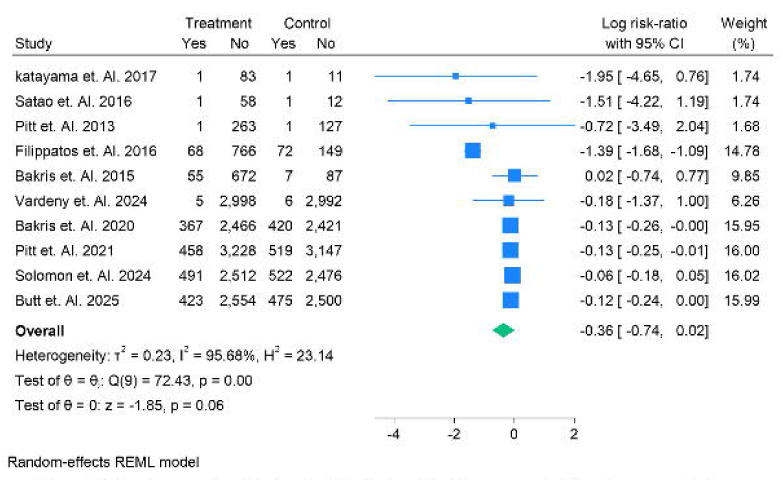
Cardiovascular Deaths in Risk Ratios for Finerenone in the given population.

### MACE

This forest plot illustrates the effect of finerenone on major adverse cardiovascular events (MACE). Small early studies (Katayama 2017, Sato 2016, Pitt 2013) showed wide confidence intervals with uncertain benefit. Filippatos 2016 suggested a favorable effect, while Bakris 2015 trended neutral. In contrast, large-scale contemporary trials (Bakris 2020, Pitt 2021, Solomon 2024, Butt 2025) consistently demonstrated modest but statistically significant reductions in MACE, with narrow confidence intervals. The pooled analysis confirmed a significant benefit (log RR –0.11 [–0.17, –0.05], p=0.00). Importantly, heterogeneity was absent (I²=0%), strengthening confidence in the robustness of this finding. Figure 4

**Figure 4.**
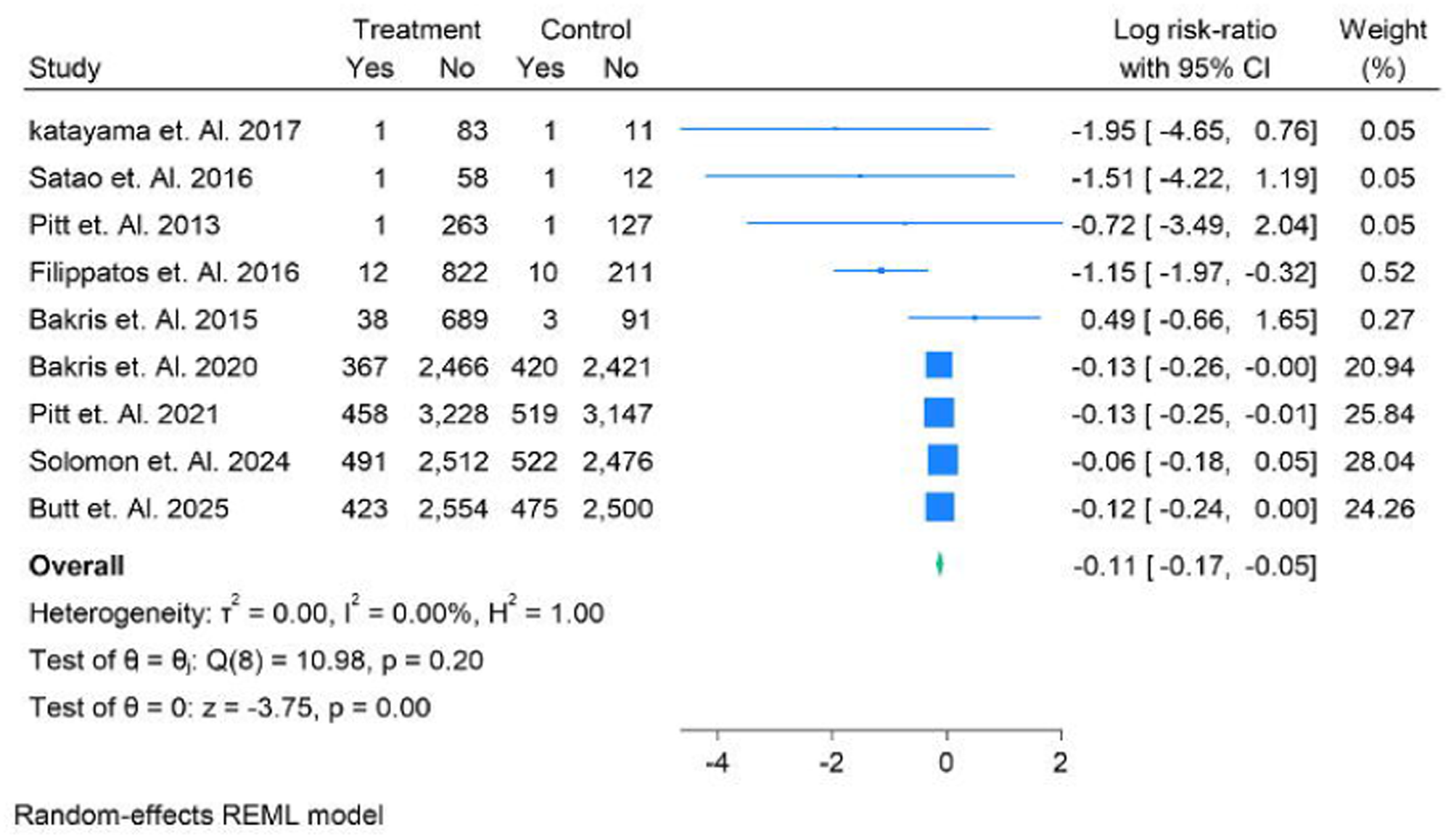
MACE in Risk Ratios for Finerenone in the given population.

### Rehospitalization

This forest plot evaluates rehospitalization outcomes with finerenone versus control. Smaller studies (Filippatos 2016, Bakris 2015) indicated significant reductions, with log risk ratios showing clear benefit. Larger contemporary RCTs produced mixed findings: Bakris 2020 showed a neutral effect, while Pitt 2021 and Butt 2025 reported modest reductions with narrower confidence intervals. The pooled analysis suggested a favorable but non-significant trend toward reduced rehospitalizations (log RR –0.66 [–1.38, 0.06], p=0.07). Heterogeneity was high (I²=96.8%), highlighting variability across trials. Collectively, results indicate potential benefit of finerenone in reducing hospitalizations, though statistical significance was not consistently achieved. Figure 5

**Figure 5.**
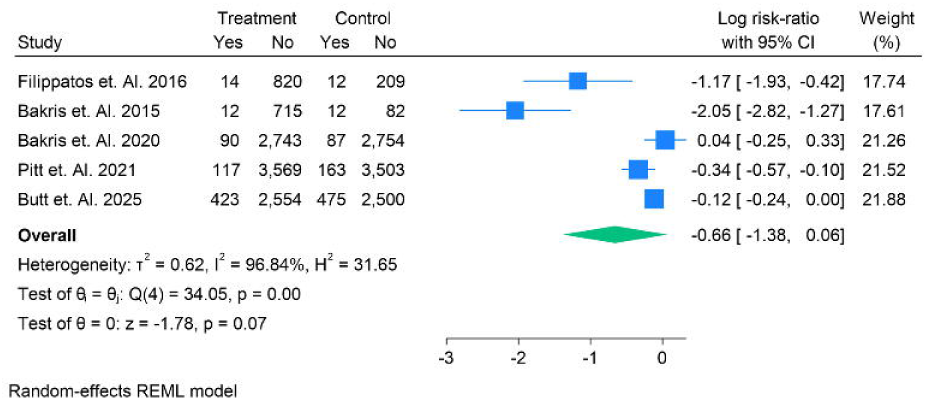
Rehospitalization during treatment in Risk Ratios for Finerenone in the given population.

### Worsening Heart Failure

This forest plot evaluates worsening renal failure adverse events with finerenone. Early small trials (Katayama 2017, Sato 2016) showed variable results, with Katayama indicating fewer events in the treatment group, while Sato was neutral. Pitt 2013 suggested increased risk, whereas Filippatos 2016 showed a modest reduction. Larger RCTs offered mixed outcomes: Bakris 2020 and Butt 2025 favored finerenone, while Pitt 2021 reported no difference. The pooled effect (log RR –0.34 [–0.90, 0.22], p=0.24) showed a non-significant trend toward fewer renal adverse events. High heterogeneity (I²=94%) highlights variability across studies, suggesting population and trial design differences influenced outcomes. Figure 6.

**Figure 6.**
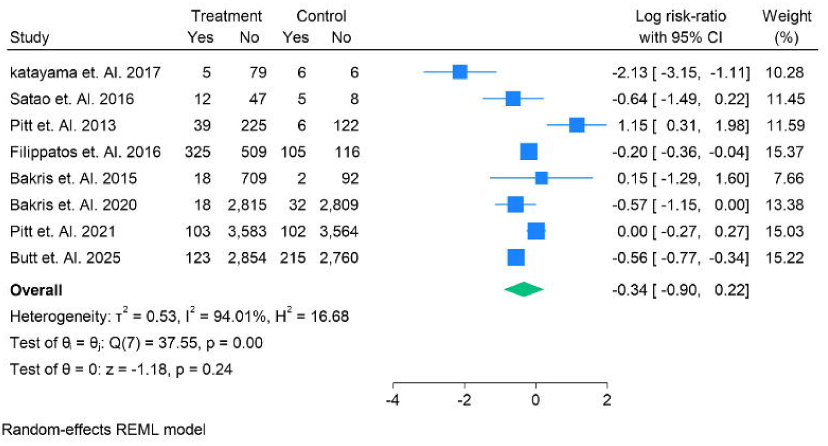
Worsening Renal Failure Adverse Events during treatment in Risk Ratios for Finerenone in the given population.

### Myocardial Infarction

This forest plot assesses myocardial infarction (MI) adverse events during finerenone therapy. Early trials (Pitt 2013, Bakris 2015) were small and imprecise, showing wide confidence intervals and no reliable conclusions. Larger contemporary studies (Bakris 2020, Pitt 2021) provided more precise estimates, both showing neutral effects with confidence intervals spanning unity. The pooled effect (log RR –0.08 [–0.28, 0.13], p=0.46) indicated no significant difference between finerenone and control. Importantly, heterogeneity was minimal (I²=1.04%), suggesting consistent findings across trials. Overall, finerenone did not significantly alter MI risk, supporting its cardiovascular safety profile without increasing ischemic events. Figure 7.

**Figure 7.**
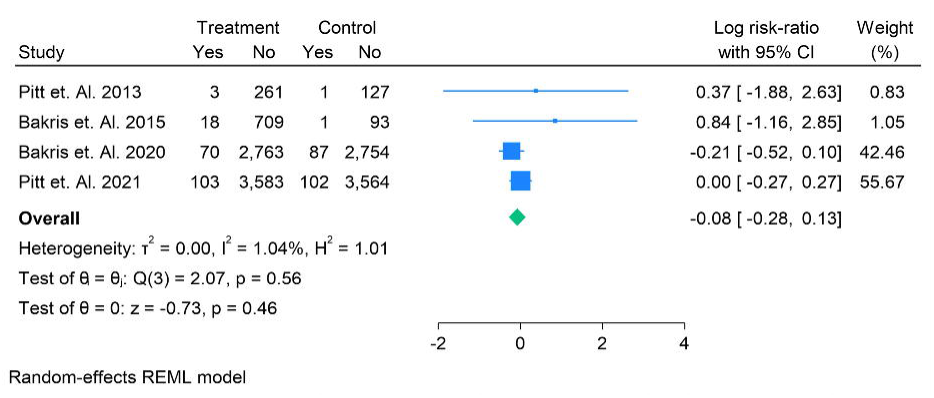
Myocardial Infarction Adverse Events during treatment in Risk Ratios for Finerenone in the given population.

### Any Adverse Events

This forest plot summarizes the incidence of any adverse events with finerenone. Small trials (Katayama 2017, Sato 2016) showed variable results, while Pitt 2013 indicated slightly higher adverse events in the treatment group. Filippatos 2016 and Bakris 2015 demonstrated neutral trends, with confidence intervals crossing unity. Large-scale trials (Bakris 2020, Pitt 2021, Solomon 2024) showed virtually identical event rates between finerenone and control arms, with extremely narrow confidence intervals. The pooled estimate (log RR 0.01 [– 0.10, 0.11], p=0.89) confirmed no significant difference, highlighting that finerenone does not increase overall adverse event risk, supporting its favorable safety profile. Figure 8.

**Figure 8.**
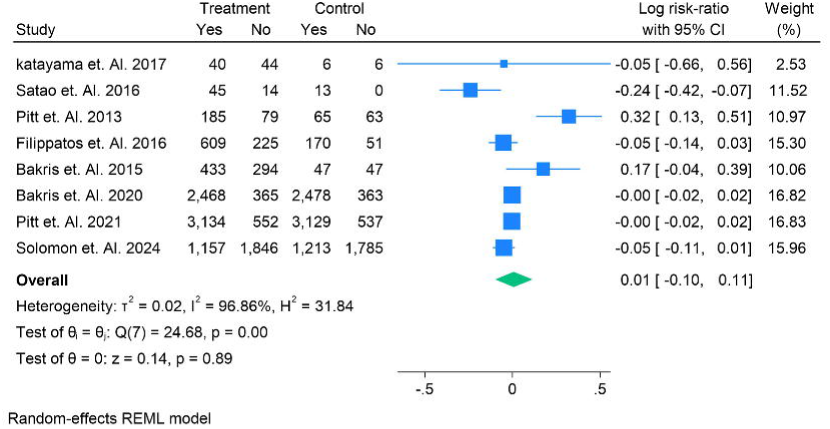
Any Adverse Events during treatment in Risk Ratios for Finerenone in the given population.

### Adverse Events

This forest plot examines adverse events leading to treatment discontinuation. Small studies (Sato 2016, Pitt 2013, Bakris 2015) showed wide confidence intervals and inconsistent results, with some suggesting higher discontinuations in the finerenone arm. Filippatos 2016 indicated fewer discontinuations with finerenone, while large-scale trials (Bakris 2020, Pitt 2021, Solomon 2024) showed near-neutral effects, with confidence intervals spanning unity. The pooled analysis (log RR –0.09 [–0.60, 0.41], p=0.72) revealed no significant difference between groups. High heterogeneity (I² =95.4%) suggests variability across populations. Overall, finerenone did not significantly increase discontinuations, supporting tolerability comparable to control. Figure 9.

**Figure 9.**
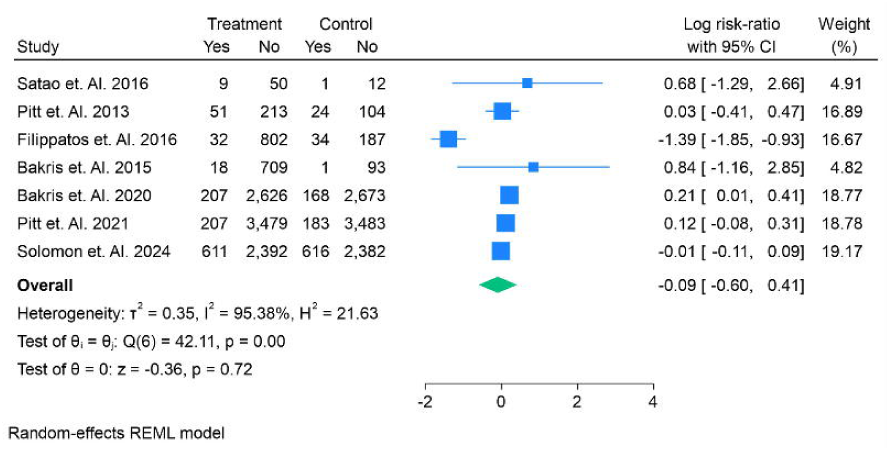
Adverse Events leading to discontinuation during treatment in Risk Ratios for Finerenone in the given population.

### Hyperkalemia

This forest plot evaluates hyperkalemia-related adverse events during finerenone therapy. Small early studies (Sato 2016, Pitt 2013) were underpowered with wide confidence intervals, showing no clear effect. Mid-sized trials (Filippatos 2016, Bakris 2015) were neutral. In contrast, large pivotal trials (Bakris 2020, Pitt 2021, Butt 2025) consistently demonstrated significantly higher hyperkalemia events in the finerenone group, with risk ratios favoring control. Solomon 2024 also showed a marked increase. The pooled estimate confirmed a significant elevation in hyperkalemia risk (log RR 0.73 [0.64–0.81], p<0.001) with no heterogeneity (I²=0%).

Thus, hyperkalemia represents a consistent safety concern with finerenone. Figure 10.

**Figure 10.**
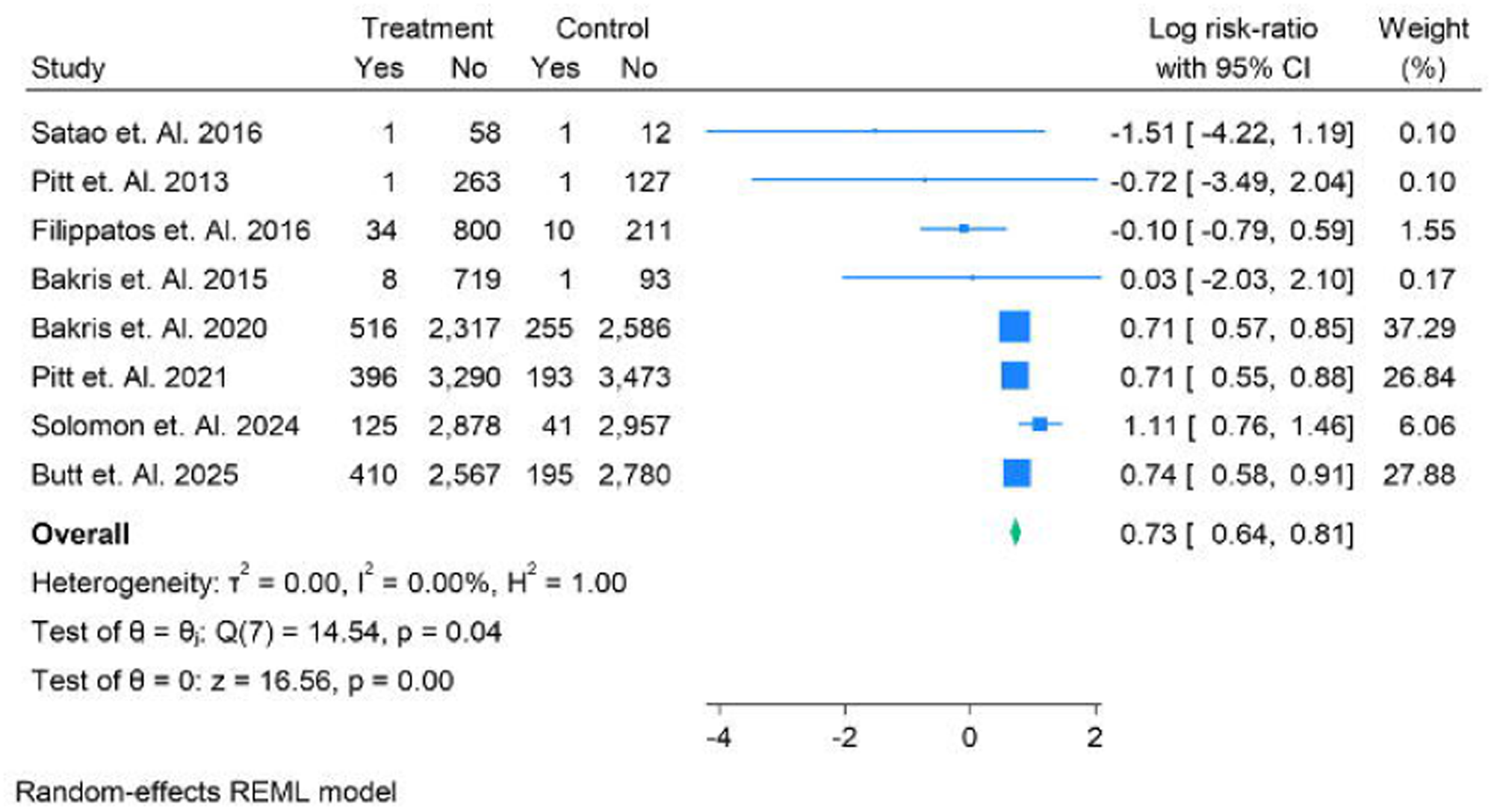
Adverse Events leading to Hyperkalemia during treatment in Risk Ratios for Finerenone in the given population.

## Discussion

Heart failure with preserved (HFpEF) and mildly reduced ejection fraction (HFmrEF) represents a growing global health burden, yet effective therapeutic strategies remain limited. In this meta-analysis of randomized controlled trials, we evaluated the efficacy and safety of finerenone in these populations. Our results indicate that finerenone provides meaningful cardiovascular benefit with an acceptable safety profile, aligning with and extending the evidence from prior meta-analyses in related populations.

The most consistent finding of our analysis was the significant reduction in **major adverse cardiovascular events (MACE)** with finerenone. This aligns with the **FIDELITY meta-analysis**, which pooled data from FIDELIO-DKD and FIGARO-DKD and reported significant reductions in composite cardiovascular outcomes, largely driven by reduced heart failure hospitalizations [28]. Our analysis extends these findings to HFpEF/HFmrEF populations, demonstrating that benefits are not restricted to patients with diabetes and CKD but also apply to broader heart failure cohorts.

While **all-cause mortality and cardiovascular death** did not reach statistical significance, both outcomes trended favorably. This is consistent with previous meta-analyses of finerenone in CKD, which also demonstrated mortality neutrality but reductions in cardiovascular morbidity. Likewise, **rehospitalization rates** showed a non-significant trend toward reduction, mirroring prior findings in CKD populations where heart failure hospitalization benefit was most pronounced [29]. Together, these findings suggest that finerenone’s greatest efficacy lies in preventing recurrent cardiovascular events and hospitalizations, outcomes that are highly relevant in HFpEF given the associated morbidity and economic impact.

Comparisons with other pharmacologic classes reinforce this interpretation. Meta-analyses of **steroidal MRAs** (spironolactone, eplerenone) in HFpEF have consistently shown modest reductions in HF hospitalizations without mortality benefit, and at the expense of higher rates of hyperkalemia and renal dysfunction. Finerenone appears to reproduce the hospitalization benefit but with **more consistent reductions in composite endpoints** and **greater tolerability**, likely attributable to its more selective non-steroidal mechanism. Similarly, metaanalyses of **SGLT2 inhibitors** in HFpEF/HFmrEF populations demonstrated robust reductions in hospitalizations with neutral mortality effects [30]. Our findings position finerenone as complementary, with SGLT2 inhibitors targeting hemodynamic and metabolic pathways while finerenone addresses inflammation and fibrosis. Together, these data support a multidrug approach to HFpEF, akin to the established “four pillars” strategy in HFrEF.

The safety profile observed in our analysis was largely reassuring. **Worsening renal function, myocardial infarction, overall adverse events, and treatment discontinuations** were not significantly different between finerenone and control groups, supporting overall tolerability. This finding is concordant with prior metaanalyses in CKD populations, which similarly reported neutral effects on kidney safety parameters when monitoring protocols were applied.

The only consistent safety signal was **hyperkalemia**, which was significantly increased in our pooled analysis, echoing results from both the FIDELITY meta-analysis and earlier pooled safety studies of finerenone. This effect is a predictable class phenomenon of MRAs and was generally manageable, with few events leading to permanent discontinuation. Compared with spironolactone, meta-analyses suggest that finerenone carries a **lower absolute risk of severe hyperkalemia**, reflecting its balanced distribution between cardiac and renal tissues. Importantly, no excess in myocardial infarction or global adverse events was observed, underscoring the cardiovascular safety of finerenone beyond the hyperkalemia concern.

## Comparison with previous meta-analyses

Our results fit into a broader pattern of meta-analytic evidence. Finerenone meta-analyses in CKD/T2DM populations demonstrated reductions in composite CV outcomes and hospitalizations, with neutral mortality and increased hyperkalemia—precisely the pattern reproduced in our HFpEF/HFmrEF-specific synthesis. Metaanalyses of spironolactone in HFpEF suggested possible hospitalization benefits but inconsistent efficacy overall, while tolerability issues limited its role. Compared with this, finerenone’s efficacy in reducing MACE appears **stronger and more consistent**, while its safety profile is **at least comparable and often more favorable**. Meta-analyses of SGLT2 inhibitors confirmed clear reductions in HF hospitalizations; finerenone complements this effect by offering additional benefit on composite CV outcomes, suggesting a role in combination therapy [31].

## Clinical implications

Taken together, the evidence suggests that finerenone should be considered a promising addition to the therapeutic armamentarium for HFpEF and HFmrEF. Its consistent effect on MACE, potential to reduce rehospitalizations, and tolerability comparable to placebo make it an attractive candidate for integration into guidelines. While hyperkalemia remains the primary limitation, this risk is predictable, monitorable, and less pronounced than with steroidal MRAs. Given the complexity of HFpEF pathophysiology, therapies with distinct mechanisms such as finerenone and SGLT2 inhibitors may prove most effective when combined, offering synergistic protection against the overlapping burdens of cardiovascular and renal dysfunction.

## Conclusion

In conclusion, this meta-analysis demonstrates that finerenone provides clinically meaningful efficacy in HFpEF/HFmrEF, particularly through consistent reductions in composite cardiovascular outcomes, while maintaining an acceptable safety profile. Our findings mirror previous meta-analyses of finerenone in CKD and extend the evidence base to non-diabetic heart failure populations. Compared with prior syntheses of steroidal MRAs and SGLT2 inhibitors, finerenone offers complementary benefit with predictable and manageable safety concerns. These results position finerenone as a credible, well-tolerated therapy with the potential to improve outcomes in HFpEF/HFmrEF, addressing a longstanding unmet need in heart failure management.

## Data Availability

supplementary file

## Authors’ contributions

All authors contributed equally to the manuscript and read and approved the final version of the manuscript.

