## supplementary file for "Efficacy and Safety of Finerenone in Heart Failure With Preserved or Mildly Reduced Ejection Fraction: A Systematic Review and Meta-Analysis of Randomized Trials"

Supplementary Files

Search String

( Finerenone[tiab] OR Kerendia[tiab] OR "BAY 94-8862"[tiab]	OR "Mineralocorticoid Receptor Antagonists"[mh]	OR (("nonsteroidal"[tiab] OR non-steroidal[tiab] OR NS-MRA*[tiab]) AND (mineralocorticoid*[tiab] OR MR*[tiab] OR "mineralocorticoid receptor antagonist*"[tiab]))) AND ("Heart Failure"[mh] OR heart failure[tiab] OR HF[tiab] OR HFrEF[tiab] OR HFpEF[tiab] OR HFmrEF[tiab] OR "systolic heart failure"[tiab] OR "diastolic heart failure"[tiab]	OR ("ejection fraction"[tiab] AND (reduced[tiab] OR preserved[tiab] OR mildly[tiab]))) AND ( efficacy[tiab] OR effectiveness[tiab] OR outcome*[tiab] OR mortality[tiab] OR "all-cause death"[tiab] OR "cardiovascular death"[tiab] OR hospitalization*[tiab] OR "composite endpoint*"[tiab]	OR "major adverse cardiovascular event*"[tiab] OR MACE[tiab]	OR safety[tiab] OR "adverse event*"[tiab] OR "drug-related side effects and adverse reactions"[mh]	OR hyperkalemia[tiab] OR "potassium"[tiab] OR "renal function"[tiab] OR eGFR[tiab] OR creatinine[tiab]	OR hypotension[tiab] OR discontinuation*[tiab] OR "serious adverse event*"[tiab] )

Table S1. Summary Findings and Demographics

| **Sr No.** | **Author Year** | **Country** | **Total Sample** | **Male** | **Female** | **Follow Up** | **Treatment Group** | **Treatment Sample** | **Control Group** | **Control Sample** | **Systolic Blood Pressure Treatment** | **Systolic Blood Pressure Control** | **BMI** | **Grade** | **Main Finding** |
| --- | --- | --- | --- | --- | --- | --- | --- | --- | --- | --- | --- | --- | --- | --- | --- |
| 1 | katayama et. Al. 2017 | Japan | 96 | 77 | 19 | 1 | Finerenone | 84 | Placebo | 12 | 137.2 ± 14.9 | 135.7 ± 16.90 | 26.68 ± 3.24 | High | **Finerenone added to RAS blockade reduced albuminuria over 90 days without hyperkalemia or renal safety signals in Japanese T2DM with diabetic nephropathy** |
| 2 | Satao et. Al. 2016 | Japan | 72 | 53 | 19 | 4 | Finerenone | 59 | Eplerenone | 13 | 112.8 ± 15.3 | 110.9 ± 13.2 | 27.8 ± 2.5 | High | In this small, randomized, double-blind Japanese study of worsening HFrEF with T2DM and/or CKD, **finerenone showed responder rates (NT-proBNP ↓ >30% by day 90) comparable to or numerically higher than eplerenone at higher doses, with a broadly similar safety profile**, supporting evaluation in larger outcomes trials. |
| 3 | Pitt et. Al. 2013 | Multicenter | 458 | 364 | 93 | 1.5 | Finerenone | 264 | Placebo | 128 | 133.8 ± 52 | 127.3 ± 38 | 28.6 ± 5.2 | High | Finerenone (BAY 94-8862) 5–10 mg/day lowered HF stress biomarkers at least as much as spironolactone but caused **less hyperkalemia and less worsening renal function** in HFrEF with moderate CKD. |
| 4 | Filippatos et. Al. 2016 | Multicenter | 1066 | 816 | 239 | 4 | Finerenone | 834 | Eplerenone | 221 | 117 ± 17 | 121 ± 19 | 29.4 ± 2.6 | High | **Finerenone lowered NT-proBNP to a similar extent as eplerenone over 90 days with comparable safety; the finerenone 10→20 mg arm showed a nominal reduction in the composite of death/CV hospitalization/emergency HF presentation vs eplerenone.** |
| 5 | Bakris et. Al. 2015 | Multicenter | 823 | 639 | 182 | 3 | Finerenone | 727 | Placebo | 94 | 137.9 ± 14.4 | 137.6 ± 15.3 | 31.75 ± 5.57 | High | Finerenone **reduced albuminuria in a clear dose-dependent manner over 90 days** without excess serious AEs; hyperkalemia-related discontinuations were infrequent. |
| 6 | Vardeny et. Al. 2024 | Multicenter | 6001 | 2732 | 3269 | 32 | Finerenone | 3003 | Placebo | 2998 | 127.6 ± 15.6 | 129.6 ± 14.2 | 27.6 ± 3.57 | High | Finerenone increased hyperkalemia risk but **maintained a lower risk** of the composite of total worsening HF events or CV death vs placebo across potassium strata with protocol-directed monitoring and dose adjustment |
| 7 | Bakris et. Al. 2020 | Multicenter | 5674 | 3983 | 1691 | 31.2 | Finerenone | 2833 | Placebo | 2841 | 138.1 ± 14.3 | 138.9 ± 14.4 | 26.79 ± 6.68 | High | **Finerenone reduced the risk of CKD progression and cardiovascular events versus placebo in T2D with CKD, with more hyperkalemia but similar overall AE rates.** |
| 8 | Pitt et. Al. 2021 | Multicenter | 7352 | 5105 | 2247 | 40.8 | Finerenone | 3686 | Placebo | 3666 | 135.8 ± 14.0 | 135.7 ± 14.0 | 135.7 ± 14.1 | High | Finerenone reduced the primary composite of CV death, nonfatal MI, nonfatal stroke, or HHF versus placebo (HR 0.87; benefit mainly from lower HHF). |
| 9 | Solomon et. Al. 2024 | Multicenter | 6001 | 3269 | 2732 | 32 | Finerenone | 3003 | Placebo | 2998 | 129.5 ± 15.3 | 129.3 ± 15.3 | 29.9 ± 6.1 | High | **Finerenone reduced the rate of total worsening heart-failure events and CV death versus placebo in HF with mildly reduced or preserved EF (rate ratio 0.84; p=0.007), with more hyperkalemia and fewer hypokalemia events.** |
| 10 | Butt et. Al. 2025 | UK | 6001 | 3241 | 2711 | 12 | Finerenone | 2977 | Placebo | 2975 | 135.9 ± 12 | 135 ± 13.5 | 32.68 ± 2.5 | High | Finerenone reduced total worsening HF events + CV death and improved symptoms **across frailty strata**, with comparable safety signals (including hyperkalemia) irrespective of frailty. |

Risk of Bias

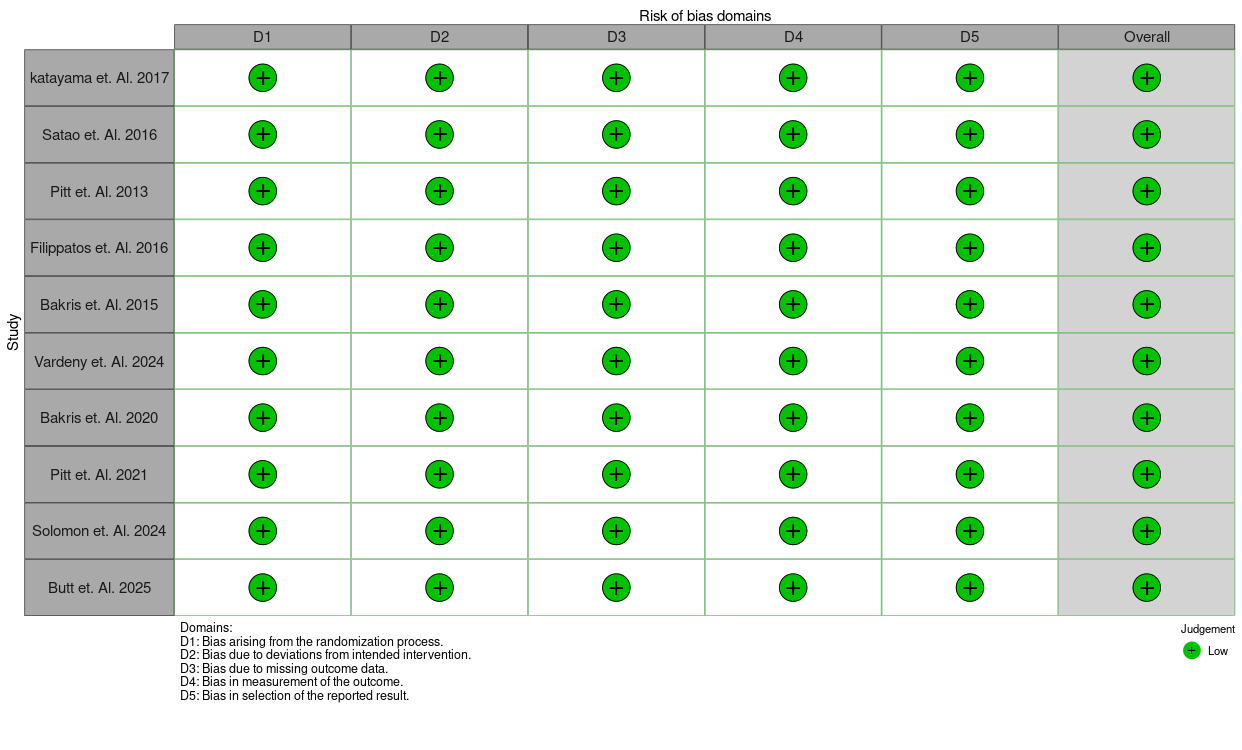
